# Follow-up study on serum cholesterol profiles in recovered COVID-19 patients

**DOI:** 10.1101/2020.12.08.20245977

**Authors:** Guiling Li, Li Du, Xiaoling Cao, Xiuqi Wei, Yao Jiang, Yuqi Lin, Vi Nguyen, Wenbin Tan, Hui Wang

## Abstract

COVID-19 patients develop hypolipidemia. However, it is unknown whether lipid levels have improved in recovered patients. In this study, a 3–6 month follow-up study was performed to examine serum levels of laboratory values in 107 discharged COVID-19 patients (mild = 59; severe/critical = 48; diagnoses on admission). 61 patients had a revisit chest CT scan. A Wilcoxon signed-rank test was used to analyze changes in laboratory values. LDL-c and HDL-c levels were significantly higher at follow-up than at admission in severe/critical cases (*p* < 0.05). LDL-c levels were significantly higher at follow-up than at admission in mild cases (*p* < 0.05). With adjustment of the factor of traditional Chinese medicine, LDL-c and HDL-c levels were significantly improved at follow-up than at admission in severe/critical cases (*p* < 0.05). Increases in HDL-c significantly correlated with increases in numbers of white blood cells (*p*<0.001) and decreases in levels of C-reactive protein (*p* < 0.05) during patients’ recovery. Residue lesions were observed in CT images in 69% (42 of 61) of follow-up patients. We concluded that improvements of LDL-c, HDL-c and incomplete absorption of lung lesions were observed at follow-up for recovered patients, indicating that a long-term recovery process could be required.

## 1. Introduction

Coronavirus disease 2019 (COVID-19), which has become a major threat to the global public health system [1], is caused by severe acute respiratory syndrome coronavirus 2 (SARS-Cov-2) [2]. As of October 3, 2020, more than 34.7 million COVID-19 infections and 1 million deaths have been reported in 188 countries and regions. China reported 90 thousands cases and 4,739 deaths which were mainly from the original epicenter, Wuhan [3]. Scientists are just beginning to understand the nature of the harm caused by this disease. The SARS-Cov-2 spike protein mediates the entrance of the virus into host cells via surface angiotensin-converting enzyme 2 (ACE2) [4, 5]. Host protease transmembrane, serine protease 2 (TMPRSS2) promotes SARS-CoV-2 entry into target cells, which are thought to be host determinants for viral infection in the initial stage [6]. COVID-19 patients may be asymptomatic or symptomatic. The time from exposure to onset of symptoms is about 5.1 days [7]. Pathologically, almost every vital organ in the body, including the lungs, heart, liver, kidneys, eyes, blood vessels, intestines, and brain, can be injured by SARS-CoV-2, leading to devastating consequences [8]. Acute and diffuse lung injuries cause increases in a series of serum cancer biomarkers in patients [9]. Damage to the lungs can be long-term for some recovered patients; this has also been observed in surviving severe acute respiratory syndrome (SARS) patients [10].

Patients with metabolic-associated preconditions are susceptible to SARS-CoV-2 attack and are likely to experience more pronounced symptoms [11]. One pathogenic cofactor associated with hypertension, obesity, diabetes mellitus, and cardiovascular disorders is hypercholesterolemia. We and others have recently reported hypolipidemia in hospitalized COVID-19 patients [12-15]. An association between decreases in lipid levels and the severity of the symptoms in patients has been revealed [12-14]. Furthermore, emerging evidence has shown that SARS-CoV-2 has a direct impact on the downregulation of lipid-metabolism-related proteins and pathways, leading to dyslipidemia [11, 16]. In addition, dyslipidemia associated with SARS-CoV-1 has also been reported [17]. Altered lipid metabolism has been shown in recovered SARS-CoV-1 patients 12 years after infection [18]. These reports demonstrate that dyslipidemia is an important clinical manifestation in patients with coronavirus-related diseases, perhaps reflecting one aspect of the complicated evolving pathological progressions seen in patients. In this study, we performed a follow-up investigation of lipid profiles and other laboratory values in COVID-19 patients from our previously reported cohort [19]. We found that there were significant improvements of coagulation and liver laboratory values, including D-dimer, antithrombin III (ATIII), fibrin degradation product (FDP), fibrinogen (FIB), alanine aminotransferase (ALT), alkaline phosphatase (ALP), and gamma-glutamyl transferase (GGT) in patients at follow-up compared to time of admission. However, partial recovery of low-density lipoprotein cholesterol (LDL-c) levels and incomplete absorption of lung lesions were observed at follow-up, indicating that a long-term recovery process and the development of sequelae such as pulmonary fibrosis could be expected in some patients.

## 2. Results

### Demographic and clinical characteristics of COVID-19 patients

A total of 107 COVID-19 cases were included in this follow-up study: 59 mild and 48 severe/critical cases (based on patient diagnosis at time of admission). A CONSORT flow diagram is shown in Figure. 1. The age for all patients was 65 (60, 70) (median (IQR)) years. The ages for patients in the two subgroups were as follows: 65 (51, 69) for mild and 66 (62, 74) for severe/critical cases (Table 1). The overall follow-up time was 100 (96, 116) days after discharge. The ratios for comorbidities, days until follow-up, and gender disparity among patients in each category are listed in Table 1. An insignificant percentage of patients in the subgroups (about 15%–18%) used cholesterol-lowering drugs (Table 1).

**Table 1.**
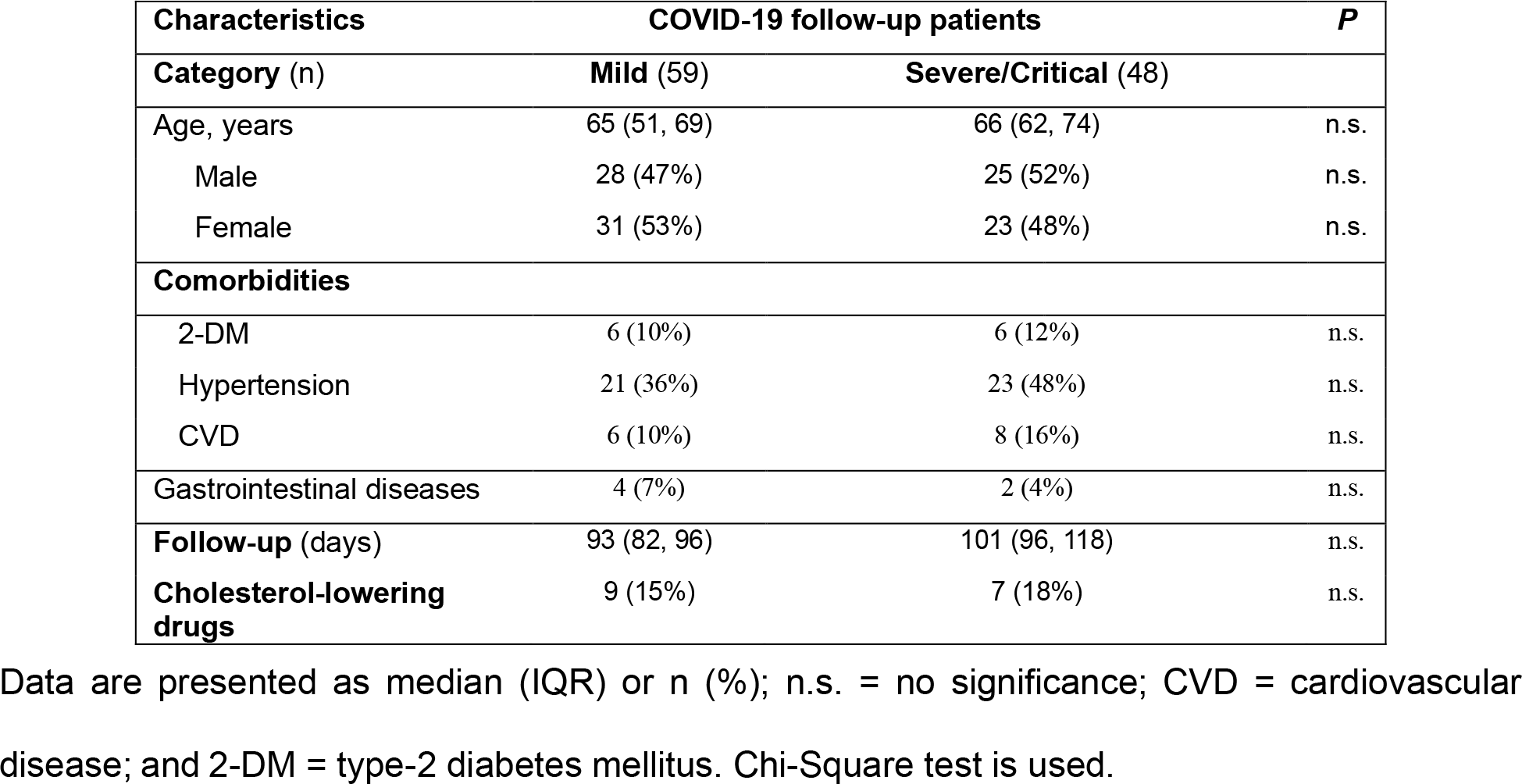
Demographic and clinical characteristics of follow-up COVID-19 patients

| Characteristics | COVID-19 follow-up patients |  | P |
| --- | --- | --- | --- |
| Category (n) | Mild (59) | Severe/Critical (48) |  |
| Age, years | 65 (51, 69) | 66 (62, 74) | n.s. |
| Male | 28 (47%) | 25 (52%) | n.s. |
| Female | 31 (53%) | 23 (48%) | n.s. |
| **Comorbidities** |  |  |  |
| 2-DM | 6 (10%) | 6 (12%) | n.s. |
| Hypertension | 21 (36%) | 23 (48%) | n.s. |
| CVD | 6 (10%) | 8 (16%) | n.s. |
| Gastrointestinal diseases | 4 (7%) | 2 (4%) | n.s. |
| Follow-up (days) | 93 (82, 96) | 101 (96, 118) | n.s. |
| Cholesterol-lowering drugs | 9 (15%) | 7 (18%) | n.s. |

Data are presented as median (IQR) or n (%); n.s. = no significance; CVD = cardiovascular disease; and 2-DM = type-2 diabetes mellitus. Chi-Square test is used.

**Figure 1.**
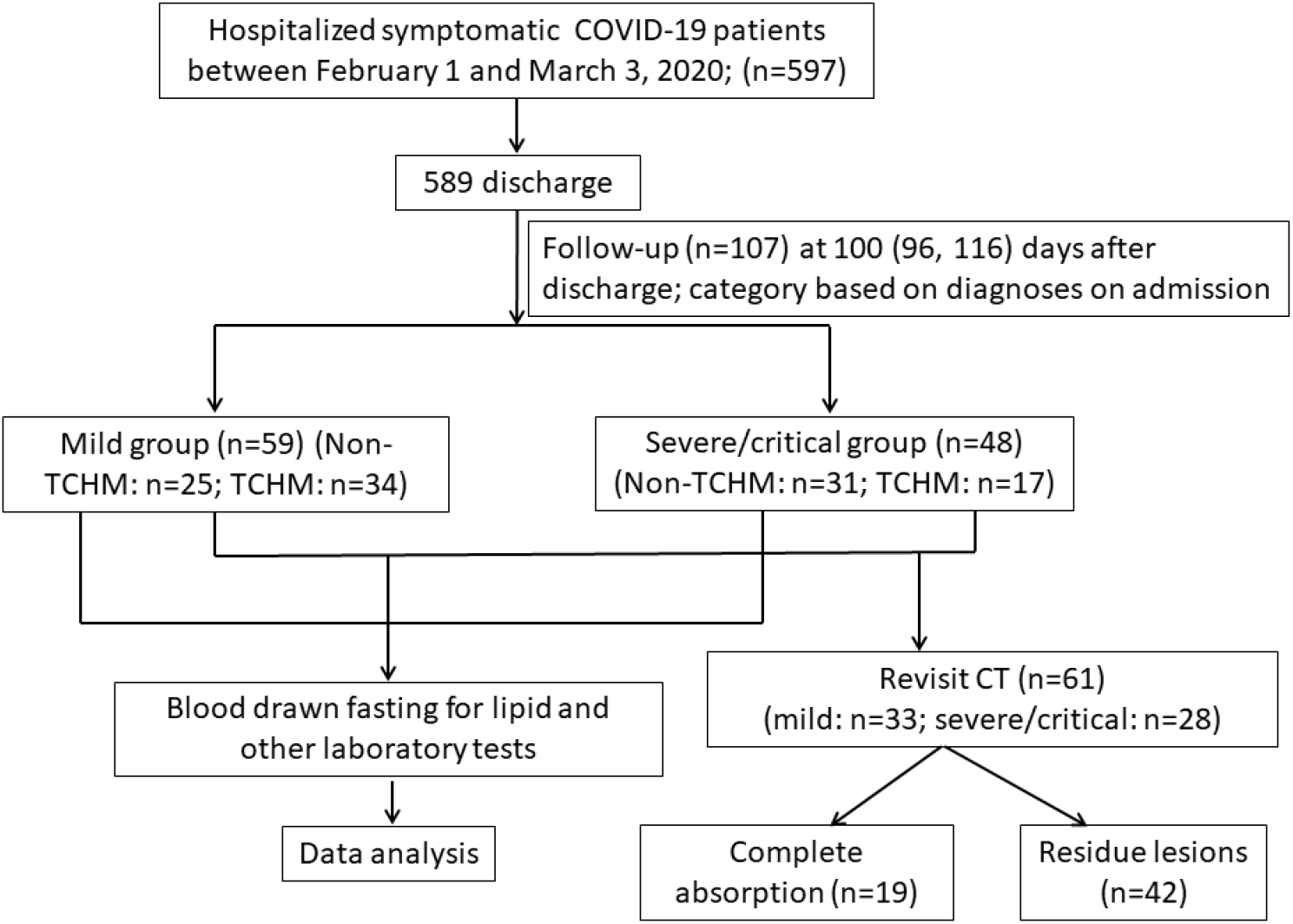
A CONSORT flow diagram for inclusion of the COVID-19 follow-up patients in this study.

### Improvements of serum cholesterol levels in recovered COVID-19 patients

Our previous studies have shown that serum cholesterol levels at the time of admission were significantly lower in COVID-19 patients than in normal subjects [12, 13]. In this follow-up study, we compared patient lipid levels at 3–6 months after discharge to those at the time of admission. Both LDL-c, HDL-c and TC were significantly higher at follow-up than at the time of admission in severe / critical cases (Table 2). LDL-c and TC levels were significantly higher at follow-up than at the time of admission in mild patients (Table 2). Surprisingly, 11% of patients (12 of 107) showed a 15% or more decrease in LDL-c levels at follow-up as compared to the time of admission; these patients included 4 mild and 8 severe/critical cases. In addition, 13% of patients (14 of 107) showed a 15% or more decrease in HDL-c levels at follow-up as compared to the time of admission; these patients included 5 mild and 9 severe/critical cases. There were two cases in which both LDL-c and HDL-c levels were 15% or more reduced at follow-up as compared to the time of admission.

**Table 2.**
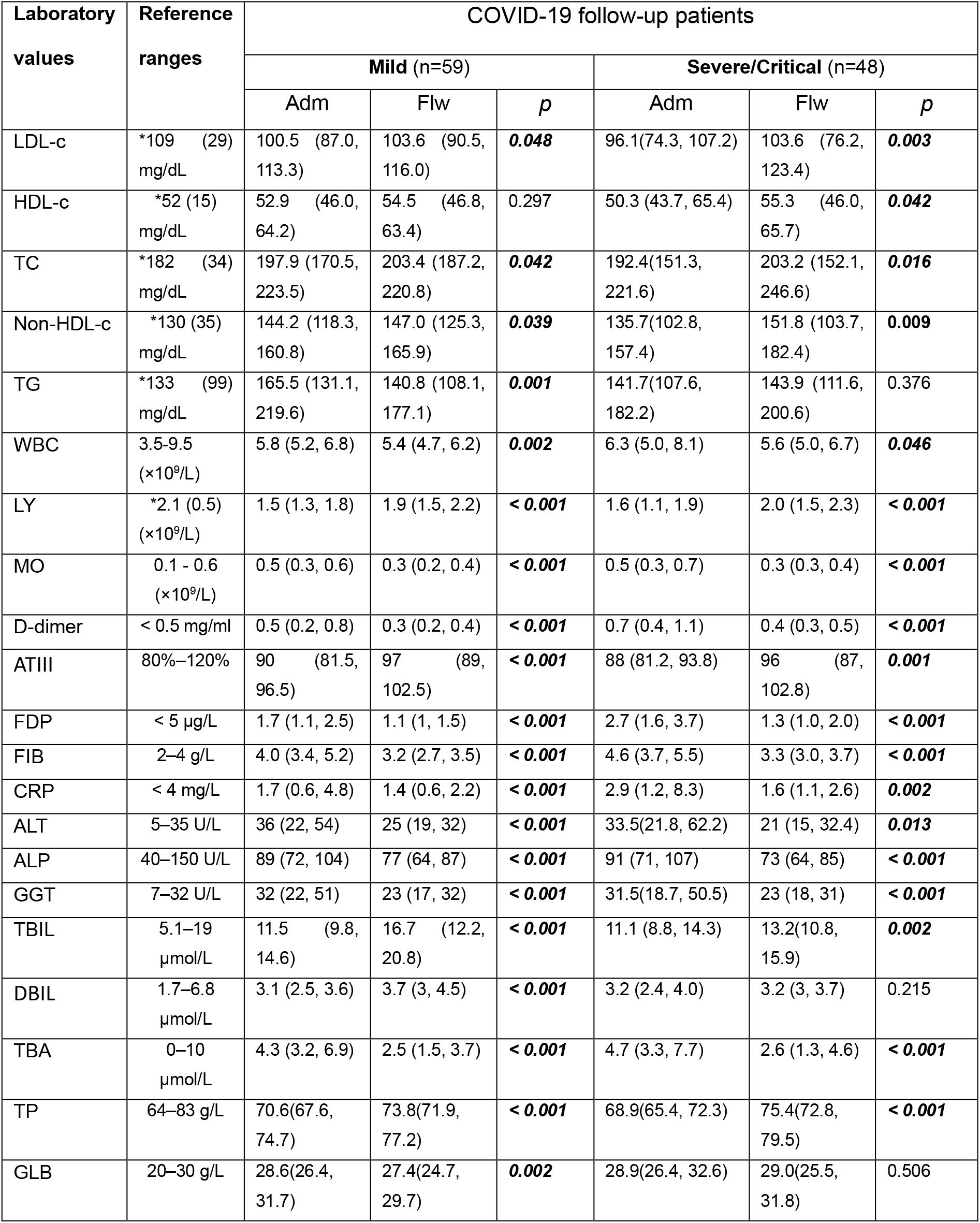
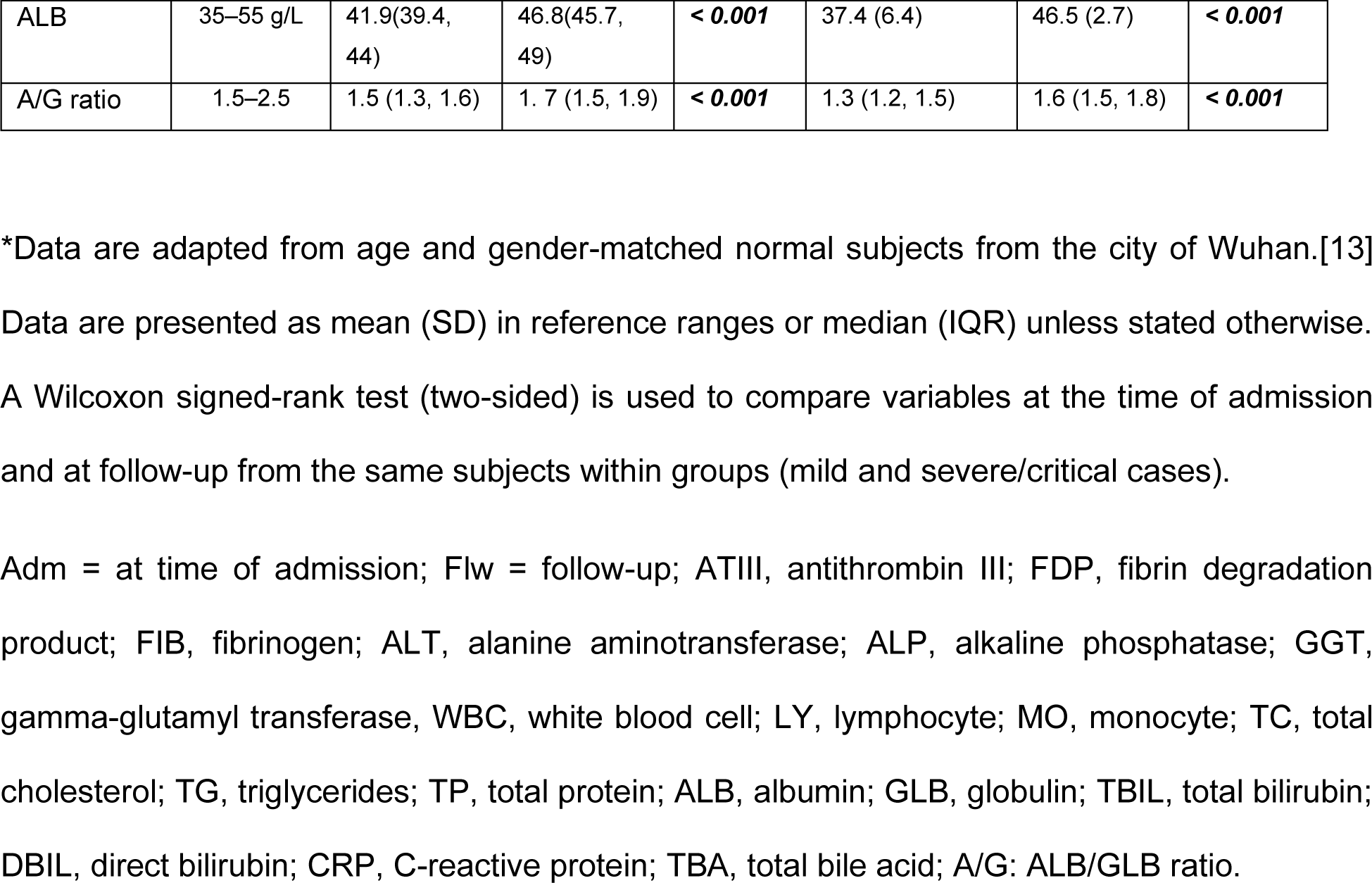
Main clinical laboratory values of COVID-19 follow-up patients

| Laboratory values | Reference ranges | COVID-19 follow-up patients |  |  |  |  |  |
| --- | --- | --- | --- | --- | --- | --- | --- |
|  |  | Mild (n=59) |  |  | Severe/Critical (n=48) |  |  |
|  |  | Adm | Flw | *p* | Adm | Flw | *p* |
| LDL-c | *109 (29)<br>mg/dL | 100.5 (87.0, 113.3) | 103.6 (90.5, 116.0) | **0.048** | 96.1(74.3, 107.2) | 103.6 (76.2, 123.4) | **0.003** |
| HDL-c | *52 (15)<br>mg/dL | 52.9 (46.0, 64.2) | 54.5 (46.8, 63.4) | 0.297 | 50.3 (43.7, 65.4) | 55.3 (46.0, 65.7) | **0.042** |
| TC | *182 (34)<br>mg/dL | 197.9 (170.5, 223.5) | 203.4 (187.2, 220.8) | **0.042** | 192.4(151.3, 221.6) | 203.2 (152.1, 246.6) | **0.016** |
| Non-HDL-c | *130 (35)<br>mg/dL | 144.2 (118.3, 160.8) | 147.0 (125.3, 165.9) | **0.039** | 135.7(102.8, 157.4) | 151.8 (103.7, 182.4) | **0.009** |
| TG | *133 (99)<br>mg/dL | 165.5 (131.1, 219.6) | 140.8 (108.1, 177.1) | **0.001** | 141.7(107.6, 182.2) | 143.9 (111.6, 200.6) | 0.376 |
| WBC | 3.5-9.5<br>(×10<sup>9</sup>/L) | 5.8 (5.2, 6.8) | 5.4 (4.7, 6.2) | **0.002** | 6.3 (5.0, 8.1) | 5.6 (5.0, 6.7) | **0.046** |
| LY | *2.1 (0.5)<br>(×10<sup>9</sup>/L) | 1.5 (1.3, 1.8) | 1.9 (1.5, 2.2) | **< 0.001** | 1.6 (1.1, 1.9) | 2.0 (1.5, 2.3) | **< 0.001** |
| MO | 0.1 - 0.6<br>(×10<sup>9</sup>/L) | 0.5 (0.3, 0.6) | 0.3 (0.2, 0.4) | **< 0.001** | 0.5 (0.3, 0.7) | 0.3 (0.3, 0.4) | **< 0.001** |
| D-dimer | < 0.5 mg/ml | 0.5 (0.2, 0.8) | 0.3 (0.2, 0.4) | **< 0.001** | 0.7 (0.4, 1.1) | 0.4 (0.3, 0.5) | **< 0.001** |
| ATIII | 80%–120% | 90 (81.5, 96.5) | 97 (89, 102.5) | **< 0.001** | 88 (81.2, 93.8) | 96 (87, 102.8) | **0.001** |
| FDP | < 5 µg/L | 1.7 (1.1, 2.5) | 1.1 (1, 1.5) | **< 0.001** | 2.7 (1.6, 3.7) | 1.3 (1.0, 2.0) | **< 0.001** |
| FIB | 2–4 g/L | 4.0 (3.4, 5.2) | 3.2 (2.7, 3.5) | **< 0.001** | 4.6 (3.7, 5.5) | 3.3 (3.0, 3.7) | **< 0.001** |
| CRP | < 4 mg/L | 1.7 (0.6, 4.8) | 1.4 (0.6, 2.2) | **< 0.001** | 2.9 (1.2, 8.3) | 1.6 (1.1, 2.6) | **0.002** |
| ALT | 5–35 U/L | 36 (22, 54) | 25 (19, 32) | **< 0.001** | 33.5(21.8, 62.2) | 21 (15, 32.4) | **0.013** |
| ALP | 40–150 U/L | 89 (72, 104) | 77 (64, 87) | **< 0.001** | 91 (71, 107) | 73 (64, 85) | **< 0.001** |
| GGT | 7–32 U/L | 32 (22, 51) | 23 (17, 32) | **< 0.001** | 31.5(18.7, 50.5) | 23 (18, 31) | **< 0.001** |
| TBIL | 5.1–19<br>µmol/L | 11.5 (9.8, 14.6) | 16.7 (12.2, 20.8) | **< 0.001** | 11.1 (8.8, 14.3) | 13.2(10.8, 15.9) | **0.002** |
| DBIL | 1.7–6.8<br>µmol/L | 3.1 (2.5, 3.6) | 3.7 (3, 4.5) | **< 0.001** | 3.2 (2.4, 4.0) | 3.2 (3, 3.7) | 0.215 |
| TBA | 0–10<br>µmol/L | 4.3 (3.2, 6.9) | 2.5 (1.5, 3.7) | **< 0.001** | 4.7 (3.3, 7.7) | 2.6 (1.3, 4.6) | **< 0.001** |
| TP | 64–83 g/L | 70.6(67.6, 74.7) | 73.8(71.9, 77.2) | **< 0.001** | 68.9(65.4, 72.3) | 75.4(72.8, 79.5) | **< 0.001** |
| GLB | 20–30 g/L | 28.6(26.4, 31.7) | 27.4(24.7, 29.7) | **0.002** | 28.9(26.4, 32.6) | 29.0(25.5, 31.8) | 0.506 |
| ALB | 35–55 g/L | 41.9(39.4, 44) | 46.8(45.7, 49) | **< 0.001** | 37.4 (6.4) | 46.5 (2.7) | **< 0.001** |
| A/G ratio | 1.5–2.5 | 1.5 (1.3, 1.6) | 1.7 (1.5, 1.9) | **< 0.001** | 1.3 (1.2, 1.5) | 1.6 (1.5, 1.8) | **< 0.001** |

\*Data are adapted from age and gender-matched normal subjects from the city of Wuhan.[13] Data are presented as mean (SD) in reference ranges or median (IQR) unless stated otherwise. A Wilcoxon signed-rank test (two-sided) is used to compare variables at the time of admission and at follow-up from the same subjects within groups (mild and severe/critical cases).
Adm = at time of admission; Flw = follow-up; ATIII, antithrombin III; FDP, fibrin degradation product; FIB, fibrinogen; ALT, alanine aminotransferase; ALP, alkaline phosphatase; GGT, gamma-glutamyl transferase, WBC, white blood cell; LY, lymphocyte; MO, monocyte; TC, total cholesterol; TG, triglycerides; TP, total protein; ALB, albumin; GLB, globulin; TBIL, total bilirubin; DBIL, direct bilirubin; CRP, C-reactive protein; TBA, total bile acid; A/G: ALB/GLB ratio.

Traditional Chinese medicine is practiced as a regular supplementary treatment to the standard care of Western medicine in China. A substantial portion of patients in this cohort (34 out of 59 in mild group and 17 out of 48 in severe/critical group) had taken either type of TCHMs at home during the onset of their symptoms or / and after discharge. In both mild and severe/critical groups, the median levels of LDL-c, HDL-c and non-HDL-c at admission were slightly but insignificantly lower in non-TCHMs subgroup as compared with the TCHM subgroup using a Mann-Whitney U test (table 3). TCHMs also did not cause any significant changes in the levels of LDL-c, HDL-c and non-HDL-c at the time of follow-up in non-TCHM subgroup as compared with TCHM subgroup (table 3, a Mann-Whitney U test). In the severe/critical group, however, LDL-c and HDL-c levels in the non-TCHM subgroup were significantly elevated at the time of follow-up as compared to admission, but not in the TCHM subgroup (Figure 2, table 3). Less patients in TCHM subgroup took cholesterol-lowering drugs than those in non-TCHM subgroup (table 3).

**Table 3.** Effects of TCHMs on lipid profiles of follow-up COVID-19 patients

| Laboratory values (mg/dL) | COVID-19 follow-up patients |  |  |  |  |  |  |  |  |  |  |  |
| --- | --- | --- | --- | --- | --- | --- | --- | --- | --- | --- | --- | --- |
|  | Mild group (59) |  |  |  |  |  | Severe/Critical group (48) |  |  |  |  |  |
|  | Non-TCHM (25) |  |  | TCHM (34) |  |  | Non-TCHM (31) |  |  | TCHM (17) |  |  |
|  | Adm | Flw | *p* | Adm | Flw | *p* | Adm | Flw | *p* | Adm | Flw | *p* |
| **LDL-c** | 95.5<br>(88.5, 116.4) | 105.2<br>(90.0, 116.6) | 0.192 | 103.3<br>(84.4, 113.9) | 101.9<br>(87.7, 118.4) | 0.139 | 87.4<br>(66.5, 105.2) | 99.0<br>(72.3, 120.6) | **0.021** | 101.3<br>(79.1, 108.7) | 112.9<br>(99.4, 129.7) | 0.089 |
| **HDL-c** | 50.7<br>(44.1, 58.7) | 52.9<br>(47.2, 61.6) | 0.142 | 57.3<br>(49.3, 65.6) | 57.7<br>(47.1, 65.5) | 0.797 | 50.3<br>(42.1, 62.6) | 57.2<br>(46.4, 66.9) | **0.002** | 51.8<br>(46.2, 72.3) | 51.0<br>(44.8, 61.7) | 0.169 |
| **Non-HDL-c** | 140.0<br>(120.6, 171.5) | 146.6<br>(126.6, 167.1) | 0.452 | 145.0<br>(114.8, 159.4) | 150.2<br>(124.2, 167.1) | **0.034** | 120.6<br>(93.2, 160.1) | 130.7<br>(95.9, 175.2) | 0.099 | 139.2<br>(114.7, 154.5) | 162.0<br>(144.2, 203.9) | 0.058 |
| **Cholesterol-lowering drugs (n)** | 8<br>(32%) | 8<br>(32%) |  | 1<br>(3%) | 1<br>(3%) | **0.003\*** | 7<br>(29%) | 7<br>(29%) |  | 0 | 0 | **0.033\*** |

TCHMs: traditional Chinese herbal medications; Adm: admission; Flw: follow-up; non-TCHM: patients did not receive any traditional Chinese herbal medications; TCHM: patients received any types of traditional Chinese herbal medications during the illness or / and recovery. Data are presented as median (IQR) or n (%). The *p* value is calculated using a Wilcoxon signed-rank test (two-sided) to compare variables at the times of admission with follow-up from the same subjects within subgroups. In both mild and severe groups, the lipid values at the time of admission or follow-up in the non-TCHM subgroup is compared with that in the TCHM subgroup using a Mann-Whitney U test; the *p* values are not shown since no statistical significance is found. \*Fisher exact test is used.

**Figure 2.**
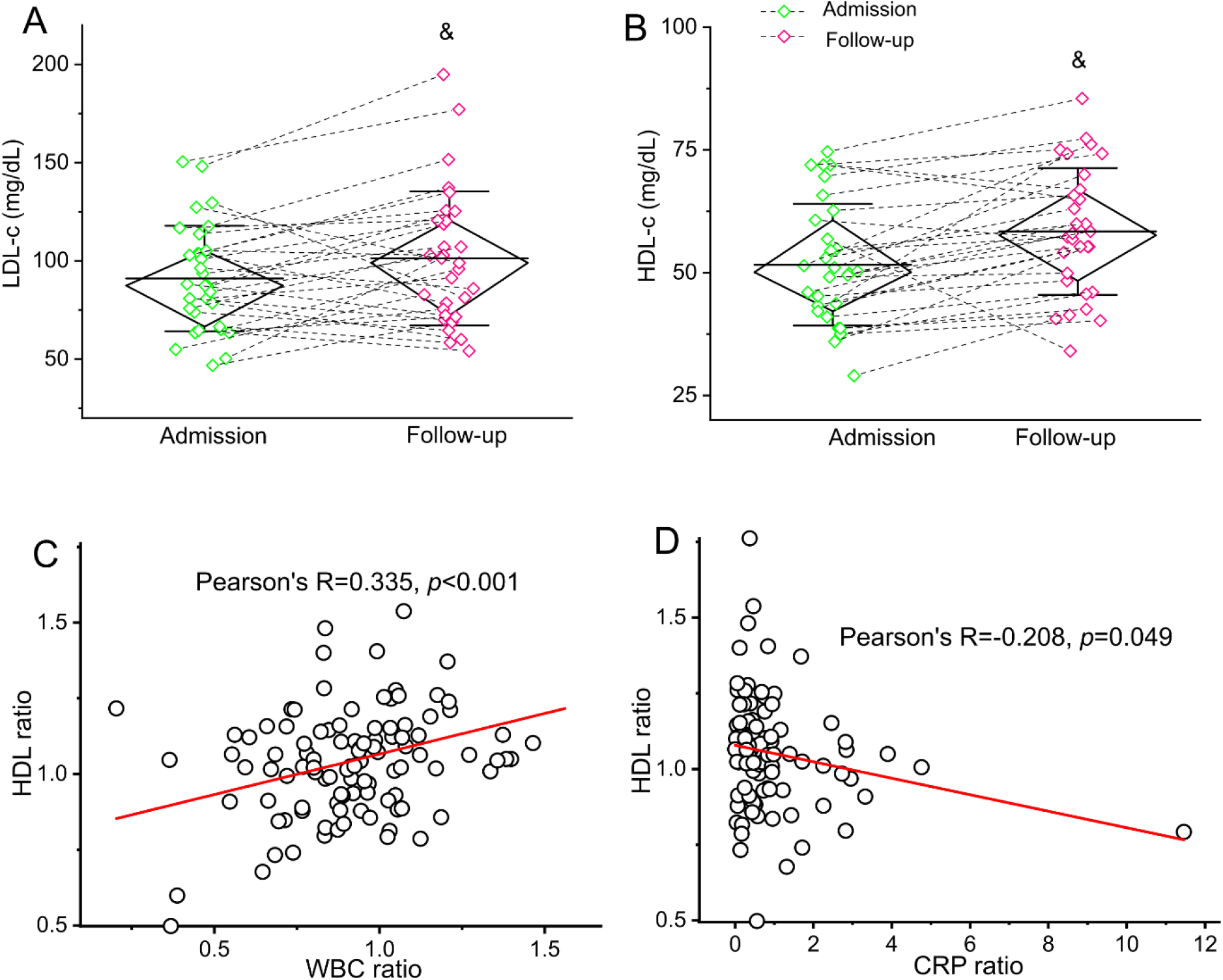
Restoration of lipid levels and correlations of ratio of WBC or CRP levels and ratio of HDL-c levels in recovered COVID-19 patients. Serum LDL-c (A) and HDL-c (B) levels are presented from COVID-19 patients in non-TCHM severe group (n=31) at follow-up and at the time of admission. Diamond boxes represent median (IQR) and whiskers represent SD in the plots. Dotted lines connect data points at the time of admission and at follow-up from the same subjects. A Wilcoxon signed-rank test is used to compare the variables at the time of admission and at follow-up from the same subjects within subgroups. Note: “&” indicates statistically significant using a Wilcoxon signed-rank test (two-sided). (C) Correlations of ratio of WBC and ratio of HDL-c levels at the time of follow-up to admission in the follow-up COVID-19 patients. Increases in HDL-c significantly correlate with increases in numbers of WBC. (D) Increases in HDL-c significantly correlate with decreases in levels of CRP at the time of follow-up to admission in the follow-up COVID-19 patients. A Pearson correlation analysis is used. non-TCHM: patients did not receive any traditional Chinese herbal medications; WBC, white blood cells; CRP, C-reactive protein.

### Improvements of other serum laboratory values in recovered COVID-19 patients

We further compared levels of other serum laboratory values at 3–6 months after discharge to those at the time of admission. Levels of numerous physiopathological markers showed significant improvements in patients at follow-up as compared to the time of admission across all subgroups; these laboratory values included LY, WBC, ALP, ALT, GGT, MO, CRP, and ALB, coagulation markers such as D-dimer, ATIII, FDP, and FIB (Table 2).

### Relationships of ratios of WBC and CRP with ratios of HDL-c

We performed a Pearson correlation analysis to calculate the correlation coefficiency between ratios of LDL-c or HDL-c and ratios of some laboratory values from follow-up to the time of admission. Increases in HDL-c significantly correlated with increases in numbers of WBC (R=0.335, *p*<0.001) and decreases in levels of CRP (R=-0.208, *p*=0.049) (Figure 2). In addition, changes in levels of LDL-c or HDL-c both significantly correlated with changes in levels of ALP (LDL-c, *R* = 0.195, *p* = 0.047; HDL-c, *R* = 0.410, *p* < 0.001), TP (LDL-c, *R* = 0.337, *p* = 0.001) and ALB (LDL-c, *R* = 0.198, *p* = 0.044; HDL-c, *R* = 0.215, *p* = 0.028).

### Incomplete absorption of lung lesions in CT findings

A total of 61 of 107 patients had a revisit CT examination at follow-up; these patients included 33 mild, 21 severe, and 7 critical cases. The main types of abnormalities in CT images include ground glass opacities (GGO), consolidation/nodules, and shadows. GGO and large diffuse shadows were two typical patterns of abnormalities found in CT images at the time of admission, suggesting diffuse and acute lung inflammations. In follow-up CT images, consolidations/nodules and small scattered shadows were the main types of residue lesions, suggesting improvements and incomplete absorbing process during recovery. A total of 31% of patients (19 of 61) showed a complete absorption of lung lesions in follow-up CT images. A total of 69% of patients (42 of 61) showed incomplete absorption, that is, residue lesions, in follow-up CT images as compared with CT findings at the time of admission.

## 3. Discussion

In this study, we performed a follow-up investigation of lipid profiles and other laboratory values on 107 recovered COVID-19 patients at 3–6 months after discharge. Our data demonstrate that levels of LDL-c and HDL-c increased significantly in severe/critical COVID-19 cases with or without adjustment of the application of traditional Chinese medicine. Coagulation and liver laboratory values, including D-dimer, ATIII, FDP, FIB, CRP, ALT, ALP, and GGT, decreased significantly across all subgroups. Furthermore, incomplete absorption of lung lesions was observed in CT images in most follow-up patients. These findings provide insight into the pathological evolution of COVID-19 during recovery and into potential long-term sequelae of the disease.

Recently, we and other investigators have reported hypolipidemia in hospitalized COVID-19 patients [12-15]. The decrease in lipid levels in patients with COVID-19 is associated with the severity of the symptoms [12-14]. These findings demonstrate that abnormalities in lipid metabolism are clinical manifestations of COVID-19 that have been underappreciated. Mild or moderate liver injuries caused by viral infection may be one important factor contributing to dyslipidemia in COVID-19 patients. Serum levels of ALT, ALP, and GGT were moderately elevated in about half of the cohort of patients in our study at the time of admission, indicating mild or moderate liver injury [13]. In this study, patient ALT, ALP, and GGT levels were significantly lower at follow-up than at the time of admission, indicating improvements in liver enzyme levels in patients during recovery. There are a couple of potential mechanisms involved in the role of cholesterol in the pathological progression of COVID-19. Wang et al. suggests that cholesterol concomitantly traffics ACE2 to viral entry sites, where SARS-CoV-2 docks in order to properly exploit entry into cells [20]. Therefore, decreased cholesterol levels in the blood may indicate severe loading of cholesterol in peripheral tissue and escalated SARS-CoV-2 infectivity.[20] Cao et al. suggests that cholesterol may facilitate an acceleration of endothelial injuries caused by SARS-CoV-2 [21]. Sorokin suggests that lowering HDL-c in COVID-19 patients may decrease the anti-inflammatory and antioxidative functions of HDL-c and contribute to pulmonary inflammation [15]. All of these hypotheses will lead to more and novel insights into the nature of this disease. The dynamics of lipid levels in a small cohort of our longitudinal study and in two cases in other reports have shown that cholesterol levels were low at the time the patients were hospitalized, remained low during disease progression, and returned to baseline levels in patients who were discharged [12, 14, 15]. To our surprise, a small portion of patients (12% to 14%) showed a decrease in LDL-c or HDL-c levels of 15% or more at follow-up as compared to the time of admission. The low lipid levels in these patients were probably due to medications or nutritional supplements taken during their own recovery process at home, for example, profound and acute dietary changes. It would be interesting to find out whether those patients with lower LDL-c or HDL-c levels at follow-up were from socioeconomically underrepresented populations. Although it is less likely, there could be a long-term sequela of lipid abnormality caused by or associated with viral infection in COVID-19 patients; there is no evidence to support the notion that SARS-CoV-2 causes long-term chronic infection.

Emerging evidence has supported coagulation as an independent mortality factor in COVID-19 patients. Coagulopathies have been found in the early stages of the disease [22-24] and in nonsurviving patients [25]. Patients have shown elevated coagulation and cardiac biomarkers such as D-dimer, fibrinogen, high-sensitivity troponin I and creatinine kinase–myocardial band [26, 27]. In our follow-up study, coagulation laboratory values, including D-dimer, ATIII, FDP, and FIB, were significantly lower in patients at follow-up as compared to the time of admission across all subgroups, indicating improvements from coagulopathies. However, we did not find significant correlations between the restoration of LDL-c or HDL-c levels and decreases in levels of these coagulation values; this suggests that recovery from dyslipidemia and improvements from coagulopathies are probably involving different pathways at different paces.

Incomplete resolution of lung lesions was observed in 69% patients in the follow-up CT examinations, suggesting pulmonary fibrosis as a potential long-term sequela for many COVID-19 patients. SARS patients have shown persistent impairment of lung function, even years after discharge [28, 29]. Pulmonary fibrosis, GGO, and pleural thickening have been reported in follow-up chest radiographs in a substantial portion of patients with Middle East respiratory syndrome coronavirus (MERS-CoV) [30]. Consistent with our findings, You et al. showed that 83.3% of COVID-19 patients had residual CT abnormalities, including GGO and pulmonary fibrosis [31]. These data suggest that aberrant wound healing in COVID-19 survivors, which is evidenced by GGO and residue lesion patterns, may lead to pulmonary fibrosis; larger studies are needed to verify this notion.

There were several limitations of this study. First, less than one-fifth of the patients from our original cohort participated in this study, which might cause a biased representative sample group from the original cohort. Second, the sample size for follow-up critical cases was limited; this might lead to an overall insignificant increase in levels of LDL-c in this subgroup. Third, many patients might have been taking various medications or remedies at home for recovery, including Chinese traditional medicines or nutritional supplements. In this study, we found that about 58% of patients in mild group and 37% of patients in severe group had taken TCHMs during their illness or / and recovery courses. Our data indicated that TCHMs might have a negative impact on the improvement of lipid profiles in patients with severe symptoms. However, due to the complexity of ingredients in those TCHMs, it will be very difficult to determine which factor(s) and how they interfere with lipid metabolisms in some patients’ recoveries in the severe group; this will need a thorough investigation in future. We, however, did not find so far that TCHMs caused any significant changes in the overall lipid profiles at the time of admission and follow-up crossing all the subgroups. Therefore, TCHMs might have a minor effect on lipid values in our patient which resulted in a negligible impact on the conclusions we drew in this study. We are aware that these data and analyses only apply to this specific Chinese population. Fourth, the lipid profiles of patients prior to discharge were crucial to determine the contributive factors to the decreased LDL-c or HDL-c levels in a small portion of patients at follow-up as compared to admission, which were lacking. Fifth, a continuous long-term follow-up is needed in order to monitor the dynamics of lipid profiles and CT abnormalities during the recovery process for a large cohort of COVID-19 patients in order to better predict potential sequelae, such as lung fibrosis; this will be our future research goal. Lastly, the characteristics of lipoproteins in our cohort was unknown. We also did not know the cellular cholesterol levels in COVID-19 in this study; such information could provide us insights into the molecular mechanisms underlying dyslipidemia in COVID-19. Whether and how cholesterol or lipoproteins participate regulation of SARS-CoV-2 entry of host cells and viral production are yet to be determined, which will our primary goal in future investigations.

Collectively, our data show improvements of LDL-c and HDL-c and incomplete absorption of lung lesions in COVID-19 patients at a 3–6-month follow-up, indicating a long-term recovery process and the development of potential sequelae such as pulmonary fibrosis.

## 4. Methods

### Study design and patients

This follow-up study was carried out at the Cancer Center at the Union Hospital of Tongji Medical College in Wuhan, P. R. China, and was approved by the Institutional Review Board (IRB) at the Union Hospital. The requirement for written informed consent was waived by the IRB committee. Our previous study included a cohort of 597 COVID-19 patients admitted to the hospital between February 1 and March 3, 2020 [13]. On June 6-10 and August 17-20, 2020, we attempted phone inquiries to all patients in this cohort; we successfully reached 260 patients and received informed consents over the phone from 144 of them to take follow-up laboratory tests. Ultimately, a total of 97 patients participated the follow-up study on June 15, 22, or 29, 2020 and 10 patients participated the follow-up study on September 3, 2020. In order to compare laboratory values at admission with those at follow-up, the patients were grouped into the same categories assigned to them in the original study, which were based on their diagnoses on admission,[13] that is, mild (n = 59) and severe/critical (n = 48) cases. Briefly, clinical diagnostic criteria and guideline were as the follows: (1) mild group, patients with onset of symptoms including fever, cough, fatigue, headache, diarrhea, and so forth, with or without mild pneumonia; (2) severe group, patients showed dyspnea, acute respiratory stress, decrease in blood oxygen saturation, lung infiltrates, multiple peripheral ground-glass opacities on both lungs; and (3) critical group, patients presented respiratory or multiple organ failure and septic shock [13]. A revisit CT scan was performed on a total of 61 patients as their standard cares of follow-up examination which was covered by their health insurance providers. A CONSORT flow diagram is shown in Fig. 1.

Many patients were prescribed traditional Chinese herbal medications (TCHM) by their local healthcare workers as a standard supplementary treatment of COVID-19 in Wuhan. Two typical TCHMs were widely given to patients with mild symptoms or at the onset of symptoms or/and after discharge from hospitals for one or two weeks according to Chinese Center for Disease Control (CDC) guidelines. The herbal ingredients for the first TCHM were as follows: bupleurum chinense, astragalus, coix lacryma-jobi, atractylodes lancea, ophiopogon japonicus, glehnia littoralis, licorice, honeysuckle, bombyx batryticatus, periostracum cicada, rhubarb, and Curcuma longa. The herbal ingredients for the second TCHM were as follows: astragalus, atractylodes macrocephala, divaricate saposhniovia root, cyrtomium fortunei, honeysuckle, eupatorium fortune, and Chenpi. In order to adjust the factor of THCM for a further analysis, we categorized patients into non-TCHM or TCHM subgroup to assess the potential effect of these THCMs on patients’ lipid profiles.

### Clinical laboratory tests and CT image acquisition

White blood cell (WBC), lymphocyte (LY), and monocyte (MO) counts were performed on a Beckman LH750 analyzer using the manufacturer’s reagents (Beckman Coulter, Brea, California, USA). A Beckman AU5800 chemistry analyzer (Beckman Coulter, Brea, CA, USA) was used for tests for the following laboratory values: ALT, ALP, GGT, LDL-c, high-density lipoprotein cholesterol (HDL-c), total cholesterol (TC), triglycerides (TG), total protein (TP), albumin (ALB), globulin (GLB), total bilirubin (TBIL), direct bilirubin (DBIL), and total bile acid (TBA). LDL-c, HDL-c, and TC were determined with standard homogeneous assays from Beckman Coulter (Catalog Nos. OSR6283, OSR6587, and OSR6616). D-dimer, ATIII, FDP, and FIB were tested on a Stago STAR analyzer using the manufacturer’s reagents (Stago, Parsippany, New Jersey, USA). C-reactive protein (CRP) was determined using a BC-5390 analyzer (Mindray, Shenzhen, Guangzhou, P. R. China). The clinical laboratory data included in this study were from blood samples drawn fasting on the morning of the day of follow-up, a standard time and procedure of blood drawn for laboratory tests in our hospital including blood sample collections at admission in our previous study.[13] Electronic data from the time of the patients’ hospitalization, including demographic information, clinical symptoms and diagnosis, laboratory tests, and treatment data, were also extracted for comparison.

CT scanners (GE LightSpeed 16, GE VCT LightSpeed 64 from GE Healthcare, Chicago, Illinois, USA) were used to take chest CT scans with the following parameters: tube voltage = 120 kVp; current intelligent control = 30–300 mA; and slice thickness/interval = 0.6–1.5 mm. The following typical abnormal patterns for viral pneumonia were reported in COVID-19 CT images:[32] ground glass opacities (GGO), consolidation/nodules, and shadows.

### Statistical analysis

Statistical analyses were performed with the statistical software SPSS (IBM, Armonk, New York, USA). Data are presented as mean (SD) or median (interquartile range, IQR). A Wilcoxon signed-rank test (two-sided) was used to compare patient laboratory values at admission and follow-up. Mann-Whitney U test was used to compare differences between two groups. A Pearson correlation analysis was used to calculate correlation coefficiency. *P* values of *p* < 0.05 were considered as statistically significant.

## Data Availability

The collection of data that supports the findings in this study is available from the Union Hospital, Wuhan, but restrictions may apply to the availability of these data, which were used under license for the current study, and so are not publicly available. Data are however available from the authors upon reasonable request and with permission of the Union Hospital, Wuhan.

## Author contributions

GL, LD, HW, and WT supervised and designed the study. GL, LD, XW, YJ, YL, and HW performed the tests and collected the data. XC, VG, and WT performed data analysis and interpretation. WT wrote the manuscript.

## Prior publication

None of the material in this manuscript has been published or is under consideration for publication elsewhere, including the Internet and conferences.

